# SUSTAINED BENEFICIAL EFFECTS OF VACCINATION ON THE CASE FATALITY RATE FOR COVID-19 INFECTIONS

**DOI:** 10.1101/2022.01.22.22269689

**Authors:** Glen H. Murata, Allison E. Murata, Douglas J. Perkins, Heather M. Campbell, Jenny T. Mao, Brent Wagner, Benjamin H. Mcmahon, Curt H. Hagedorn

## Abstract

**Objective:** To evaluate the benefits of vaccination on the case fatality rate (CFR) for COVID-19 infections.

**Design:** Multivariate modeling of data from electronic medical records

**Setting:** 130 medical centers of the United States Department of Veterans Affairs

**Participants:** 339,772 patients with COVID-19 confirmed by nucleic acid amplification testing as of September 30, 2021

**Methods:** The primary outcome was death within 60 days of the diagnosis. Patients were considered vaccinated if they had completed a full series >= 14 days prior to diagnosis. Cases presenting in July - September of 2021 were considered to have the delta variant. Logistic regression was used to derive adjusted odds ratios (OR) for vaccination and infection with delta versus earlier variants. Models were adjusted for demographic traits, standard comorbidity indices, selected clinical terms, and 3 novel parameters representing all prior diagnoses, all prior vital signs/ baseline laboratory tests, and current outpatient treatment. Patients with a delta infection were divided into 8 cohorts based upon the time from vaccination to diagnosis (in 4-week blocks). A common model was used to estimate the odds of death associated with vaccination for each cohort relative that of all unvaccinated patients.

**Results:** 9.1% of subjects had been fully vaccinated, and 21.5% were presumed to have the delta variant. 18,120 patients (5.33%) died within 60 days of their diagnoses. The adjusted OR for delta infection was 1.87 +/- 0.05 which corresponds to a relative risk of 1.78. The overall adjusted OR for prior vaccination was 0.280 +/- 0.011 corresponding to a relative risk of 0.291. The study of vaccine cohorts with a delta infection showed that the raw CFR rose steadily after 10-14 weeks. However, the OR for vaccination remained stable for 10-34 weeks.

**Conclusions:** Our study confirms that delta is substantially more lethal than earlier variants and that vaccination is an effective means of preventing COVID death. After adjusting for major selection biases, we found no evidence that the benefits of vaccination on CFR declined over 34 weeks.

## INTRODUCTION

Recent studies have shown an alarming decrease in the effectiveness of COVID vaccines over time (1-3). The metrics for effectiveness included infection and mortality rates (4, 5). It is imperative that we understand the mechanisms by which vaccines begin to fail. Such efforts can be facilitated by a robust framework for classifying vaccine effects. The most straightforward method is to use a probabilistic approach. Patients who succumb to a contagious disease must first be exposed, then develop an infection as a result of the exposure, and then die as a consequence of the infection. The risk of death in a population observed for a given time is thus the joint probability of these 3 events or P(exposure, infection, death). From the chain rule of probability theory:

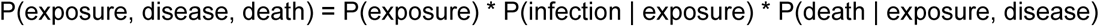

Likewise, the risk of infection is the joint probability of the first 2:

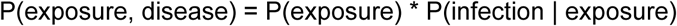

It is important to separate these metrics into their underlying risks because the latter represent separate targets for interventions. For example, COVID precautions focus on P(exposure), while anti-virals target P(death | exposure, disease). Vaccination has favorable effects on P(infection | exposure) and P(death | exposure, disease) by promoting an immune response. It will hopefully decrease P(exposure) once herd immunity is achieved. Thus, vaccine effectiveness might vary depending upon which risk is targeted. Moreover, changes in one risk may be offset by changes in another - leading to erroneous conclusions about vaccine effectiveness. For example, beneficial effects of the vaccine on P(infection | exposure) may be diminished by abandoning COVID precautions which increases P(exposure).

One problem with certain models for vaccine effectiveness is that they do not account for the risk of exposure. Doing so requires adjustments for patient behaviors and community level effects. The former include adherence to COVID precautions such as masking, social distancing, handwashing, avoidance of large crowds, testing of contacts, and working from home. The latter include the prevalence of the virus, its infectivity, the extent to which the community embraces COVID precautions, and government mandates. As a result, increases in infection and mortality rates may be related to diminished vaccine effectiveness, changes in exposures, or both. In this study, we focused on the third risk or P(death | exposure, disease). This term is analogous to the case fatality rate (CFR). We chose this outcome to assess vaccine effectiveness because, unlike infection and mortality rates, it is not affected by unmeasured patient behaviors and environmental factors.

Measuring effectiveness in observational studies requires a robust approach to confounding because treatment is not randomly allocated across a population. Confounding is introduced when the treatment (vaccination) affects the outcome, the condition for which treatment is indicated (a pre-existing condition) affects the outcome, and there is an association between the severity of the condition and the likelihood of treatment. In most cases, the benefits of treatment are offset by their preferential use in patients with a poorer prognosis. As such, the benefit can only be revealed when the bias is removed by multivariate analysis which separates the independent effects of treatment and associated comorbidities. The challenge is that there are hundreds of conditions that may serve as confounders. Co-morbidity scores may not be suitable for this purpose because they do not represent all conditions that pose a risk. Critical findings on vital signs and laboratory tests may also serve as confounders. The most robust solution is to do a systematic survey of all high-risk conditions from several domains in the medical record and adjust the effect of vaccination by some aggregate measure of their effect. We developed and applied such procedures for individual ICD10 codes, vital signs, commonly used laboratory tests, and outpatient medications in this study.

Finally, patients can have certain traits that have a major effect on prognosis but are not easily measured or well represented by their underlying diagnoses. For example, nursing home patients have a poor prognosis but are not easily identified if such care is delivered through a nursing home contract or private arrangement. Nevertheless, it is important to control for these confounders to get an unbiased estimate of vaccine effect. We felt that the timing of vaccination might be used as a proxy for these traits because VA prioritized its delivery of the vaccine. The COVID-19 Vaccination Plan for the Veterans Health Administration acknowledged that elderly patients, certain ethnic groups, and those with major comorbidities were at high risk of death or complications. It also authorized a population-based risk stratification plan for vaccine administration and its implementation when supplies were limited. For this reason, we examined vaccine effect for cohorts defined by the time from vaccination to diagnosis. Members of cohort had the same priority for vaccination – removing the criteria as potential confounders. Multivariate analysis within each cohort was then used to adjust the effect of vaccination for several patient covariates. This dual approach to confounders also reduces the bias resulting from patient self-selection – that is, patients seeking early vaccination if they believed their health was poor or deferring vaccination if they believed their health was good. Because the estimates were unbiased, it was possible to compare vaccine effectiveness across the cohorts to determine whether it declined over time. Our choice of endpoints, a more robust approach to measured confounders, and a stratified analysis to handle the urgency of vaccination provided new insights into the benefits of vaccination on CFR.

## METHODS

Cases were identified through the COVID Shared Data Resource (CSDR) of the US Department of Veterans Affairs (VA). CSDR contains cases reported by 130 medical centers and may include non-veterans referred to VA by other agencies. Membership in this registry requires at least one positive nucleic acid amplification test. Subjects were included in this study if their index infections occurred before October 2021. Delta variants of the virus were considered the infecting agents for those presenting in July, August, or September of 2021. Although sporadic cases of delta were reported in late May, delta was the predominant variant over the time we selected. The primary outcome was death within 60 days of the diagnosis. The outcome was retrieved from the CSDR, which assigns a 1 to those who died and 0 otherwise. The cohort was followed through November 2021 so that each subject reached a definitive endpoint.

VA maintains two databases containing information on COVID vaccination. CSDR has a robust and highly vetted registry of patients who have been vaccinated within and outside of the agency. The immunization domain of the Corporate Data Warehouse (CDW) contains similar information but is less structured and contains duplicates. The CDW data were scrubbed and re-organized to match the CSDR format. Cases identified in CDW, but not in CSDR, were added to the latter to create a pooled vaccine registry. Patients were considered vaccinated if they had received 1 dose of the Johnson & Johnson product or 2 doses of any other formulation at least 14 days prior to the diagnosis of COVID.

PDeathDx refers to the predicted probability of death based upon 153 ICD10 category diagnoses (6). Pre-existing conditions were identified by reviewing all diagnoses entered into the electronic medical record during outpatient visits, as updates to the patient problem list, or at the time of hospital discharge. “Pre-existing” refers to entries made up to 14 days prior to the COVID diagnosis. ICD9 codes were converted to ICD10 using a crosswalk provided by the Centers for Medicare/ Medicaid Services. A “category diagnosis” was defined as all characters preceding the decimal point for ICD10 codes or the ICD9 equivalent. Each patient was deemed to have (or not have) each category diagnosis prior to COVID. A proprietary computer program was used to identify all patients with a given condition who died or survived, as well as all patients without that condition who died or survived. The software used these cell frequencies to derive the relative risk (RR) of death associated with the condition along with the confidence interval (CI). CIs were adjusted for multiple comparisons by the Bonferroni method. A category diagnosis was considered to have a significant effect on the outcome if the lower limit for the CI was ≥ 1.5 or the upper limit for the CI was ≤ 0.80. The procedure was thus used to identify conditions that were either high-risk or protective. Stepwise logistic regression identified those diagnoses that were independent predictors of death. The model was then used to generate a predicted probability of death (PDeathDx) for each subject.

PDeathLabs refers to the predicted probability of death based upon 49 parameters derived from complete value sets for 4 vital signs (systolic blood pressure, diastolic blood pressure, O2 saturation, and body mass index) and 7 routine laboratory tests (estimated glomerular filtration rate, ALT, hematocrit, serum albumin, low-density lipoprotein cholesterol, high-density lipoprotein cholesterol, and hemoglobin A1c). Entries for these 11 clinical measurements were retrieved if their recorded dates were ≥ 14 days prior to the diagnosis of COVID. 13 parameters were derived for each type of measurement to reflect criteria used by practitioners to assess metabolic control (total = 13 * 11 = 143). Logistic modeling showed that 49 of these parameters were independently predictive of death (7). The model was used to assign a predicted probability of death (PDeathLabs) based on these clinical measurements to each subject.

Current treatment was identified by reviewing all outpatient medications active on the 14^th^ day prior to the COVID diagnosis. A patient was considered on treatment if (s)he still had a supply of medications from their most recent “fill” on the cutoff date. The VA system assigns each formulation to one or more drug classes. A process identical to the one above was used to assign a RR and CI to each of the 343 VA drug classes. AggRiskRx refers to the protective effect of 8 VA drug classes with an upper boundary for CI ≤ 0.80. This definition presumes that a protective effect goes beyond neutralizing the underlying condition and is therefore likely to be independent of its initial indication. An aggregate effect for all 8 classes was derived by log transforming the relative risk for each and adding the transformed values. This approach assumes that their effect was independent and that the aggregate effect was the product of the individual RR. We did not examine high-risk drugs because the RR of pre-existing conditions reflects the underlying disease as well as the drugs used to treat the condition.

Age at diagnosis, gender, self-reported race and ethnicity, veteran status, smoking history and use of supplemental oxygen were retrieved from the CSDR. The CSDR was also interrogated for the 2-year (Charl2Yrs) and lifetime Charlson Comorbidity Index (CharlEver) and the 2-year (Elix2Yrs) and lifetime Elixhauser (ElixEver) scores.

### Statistical methods

Univariate analysis was used to compare the attributes of patients who died and survived. Group differences in nominal variables were tested by chi-square analysis. Group differences in continuous variables were examined by the student’s t-test or Mann-Whitney U-test.

#### Main model

Stepwise logistic regression was used to construct a multivariate model for COVID death in the entire sample. The dependent variable was death within 60 days of the diagnosis. The predictor of interest was prior vaccination for COVID-19. Covariates included age, gender, race, ethnicity, veteran status, current smoking, use of supplemental oxygen, PDeathDx, PDeathLabs, AggRiskRx, Charl2Yrs, CharlEver, Elix2Yrs, ElixEver and infection with the delta versus earlier variants. Variables were entered in a stepwise fashion with a P-to-enter of 0.01 and to remove of 0.05. The model was used to derive an overall predicted probability of death (PDeath) for each patient. The ability of PDeath to discriminate between the two groups was assessed by the area under its receiver operator characteristic (ROC) curve. An adjusted odds ratio (OR) and its 95% CI was derived for the vaccination term. A standard on-line calculator was used to convert the adjusted OR to an equivalent RR. The identical procedure was used to evaluate the delta term.

#### Early variants versus delta

Separate models were developed for early variants (pre-July) and for delta (July - September) using the methods described above. The objective was to determine if predictors of death had changed significantly and if the effectiveness of prior vaccination differed for the two groups.

#### Vaccine cohorts

Eight patient cohorts infected with delta were assembled based upon the time from vaccination to the date of diagnosis (VxToDx). Each cohort was comprised of patients whose VxToDx fell within a 4-week interval. Cohort 1 was vaccinated ≥ 2 and < 6 weeks prior to diagnosis, cohort 2 (≥6 and <10 weeks), cohort 3 (≥10 and <14 weeks), cohort 4 (≥14 and <18 weeks), cohort 5 (≥18 and <22 weeks), cohort 6 (≥22 and <26 weeks), cohort 7 (≥26 and <30 weeks), and cohort 8 (≥30 and <34 weeks). Patients vaccinated ≥ 34 weeks prior to diagnosis were excluded because the cohorts were small. CFR was calculated for each cohort and plotted against cohort number (**Figure 1**).

**FIGURE 1:**
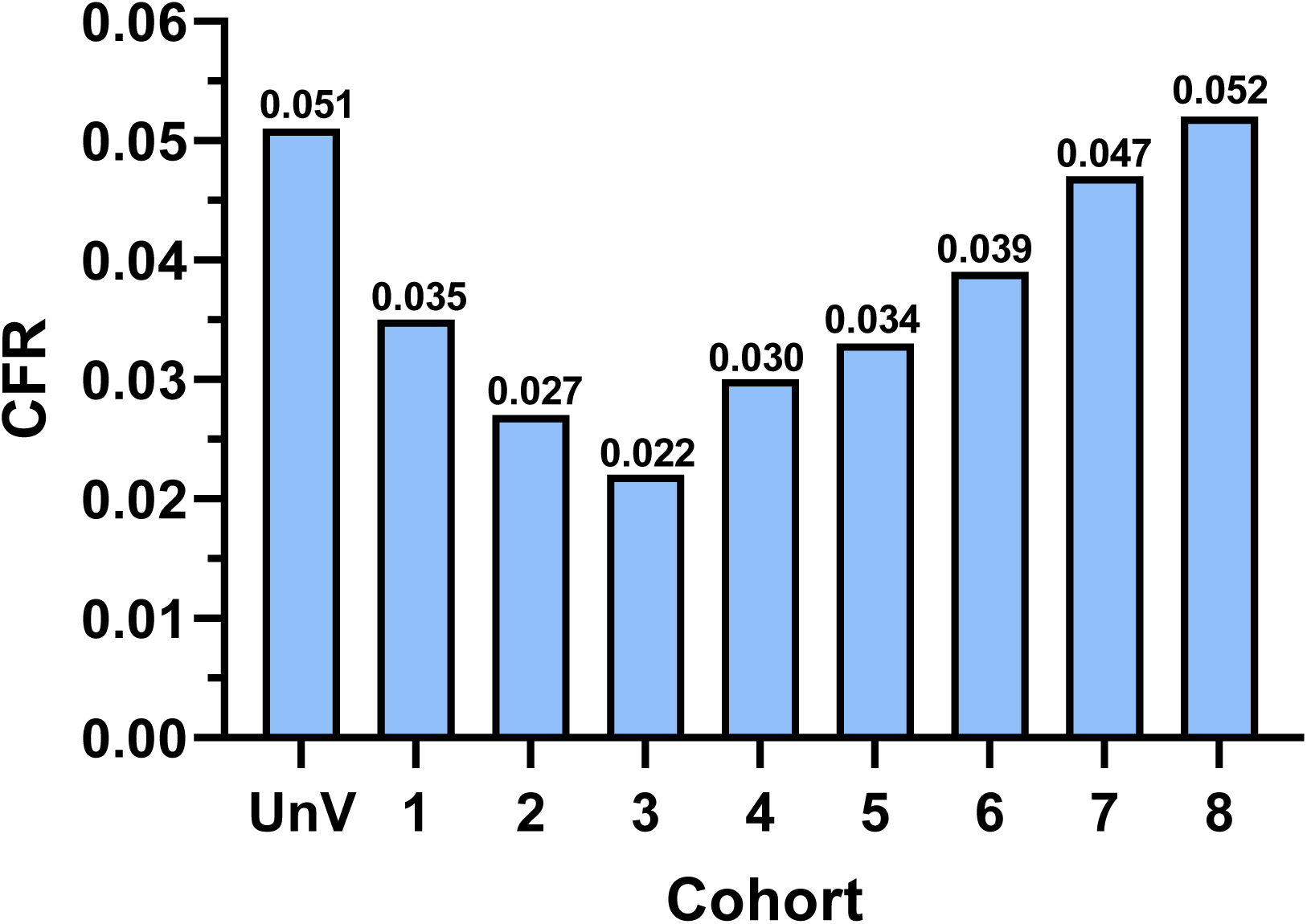
Unadjusted case fatality rates (CFR) for delta infections by time from vaccination to diagnosis (in 4-week blocks). The CFR for unvaccinated (UnV) patients was 5.06%. Cohort 1 was vaccinated ≥ 2 and < 6 weeks prior to diagnosis, cohort 2 (≥6 and <10 weeks), cohort 3 (≥10 and <14 weeks), cohort 4 (≥14 and <18 weeks), cohort 5 (≥18 and <22 weeks), cohort 6 (≥22 and <26 weeks), cohort 7 (≥26 and <30 weeks), and cohort 8 (≥30 and <34 weeks). The CFR is shown above each column for the unvaccinated individuals and the 8 cohorts. Note that CFR reached a nadir for cohort 3 and rose monotonically across cohorts 3 to 8.

Logistic modeling was used to derive cohort-specific adjusted odds ratios for vaccination. For cohort 1, patients in vaccine cohorts 2-8 were excluded from the data set of all delta patients. A logistic model was then fitted to the remaining cases. This model was comprised of age at diagnosis, male gender, use of supplemental oxygen, current smoking, prior vaccination, PDeathDx, PDeathLabs, Charl2Yrs, and CharlEver. This model was chosen because preliminary regressions showed that all variables were significant predictors of death for every cohort. The subset models also had similar power to discriminate between non-survivors and survivors (ROC areas from 0.810 to 0.816). Vaccine effect was therefore adjusted for 8 other demographic and clinical variables and expressed as the odds of death for the cohort relative to that of all unvaccinated patients. This process was repeated for the remaining cohorts. The adjusted odds ratios for vaccination were plotted against cohort number (**Figure 2**).

**FIGURE 2:**
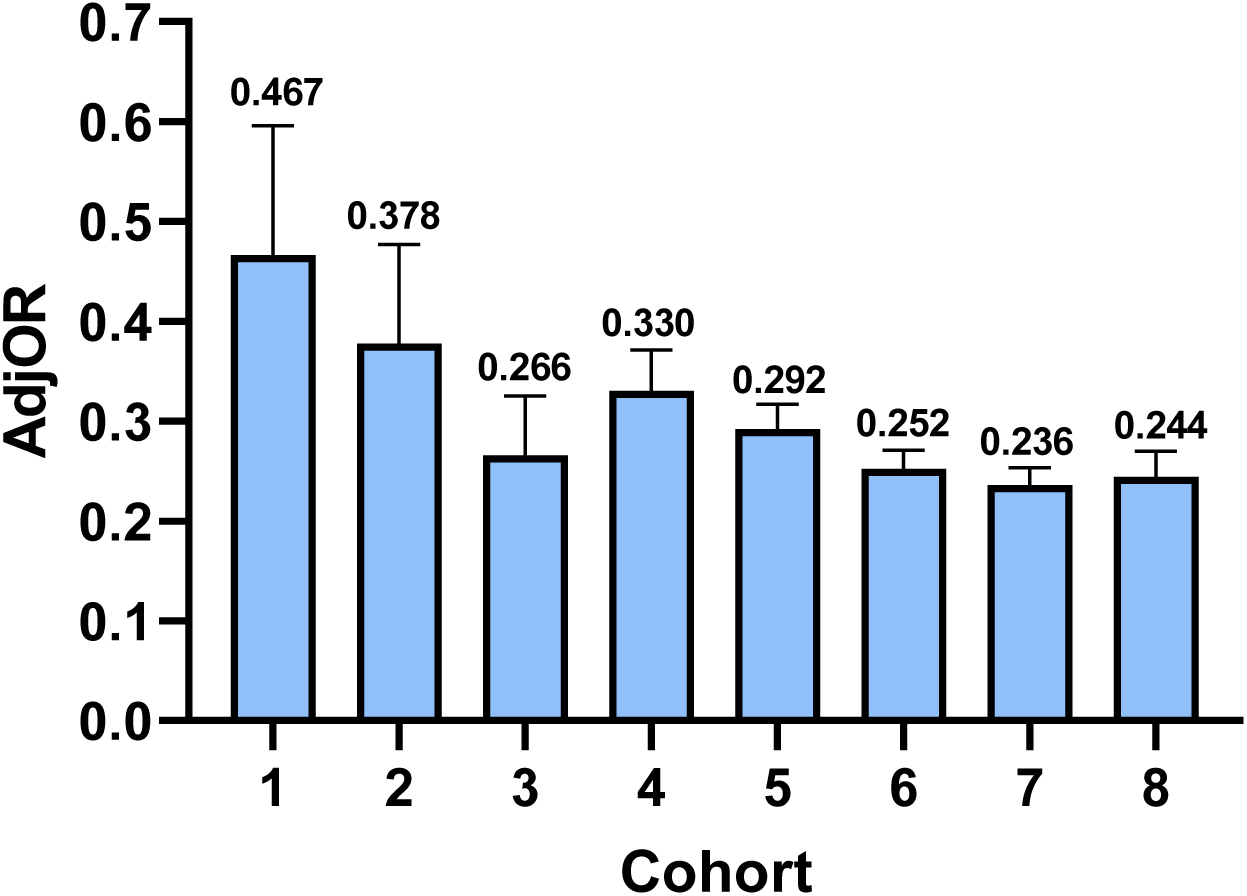
Adjusted odds ratios for vaccination (AdjOR) by time from vaccination to diagnosis (in 4-week blocks). Cohorts are defined in the caption for **Figure 1**. A cohort specific AdjOR is the odds of death for that cohort relative to that of all unvaccinated patients. The AdjOR is shown above the standard error of the OR for the 8 cohorts. Note that the benefit of vaccination for preventing COVID death remained relatively stable across cohorts 3 to 8 (10-34 weeks).

## RESULTS

On September 30, 2021, there were 347,220 COVID patients in VA’s COVID Shared Data Resource. 339,772 (or 97.9%) had at least one pre-existing condition and form the basis for this report. The mean age at the time of diagnosis was 58.6 ± 16.7 years; 84.1% were male; 22.9% were members of a racial minority; 9.0% were Hispanic; 95.8% were veterans; 0.7% were on supplemental oxygen; and 11.8% were current smokers. 9.1% had been fully vaccinated at least 14 days prior to the COVID diagnosis. The median interval between vaccination and diagnosis was 154 days (interquartile range 111 to 185). 21.5% acquired their infections after July 1, 2021 and were presumed to have the delta variant. Overall, 18,120 patients (5.33%) died within 60 days of their diagnosis.

**Table 1** shows the results of univariate analysis comparing non-survivors and survivors. Non-survivors were older and more likely to be male, white, and on supplemental oxygen but less likely to be Hispanic or current smokers. Vaccinated patients were less likely to die than the unvaccinated (3.95% vs 5.47%, respectively; P < 0.001). The case fatality rate was lower for those acquiring delta than earlier variants (4.64% vs 5.52%; P < 0.001). This finding persisted even when vaccinated patients were removed from the analysis (5.06% vs 5.55%; P < 0.001).

**TABLE 1:** CHARACTERISTICS OF NON-SURVIVORS AND SURVIVORS.

| Attribute | Non-Survivors | Survivors | P_Value |
| --- | --- | --- | --- |
| Age at diagnosis (years) | 76.1 ± 11.2 | 57.6 ± 16.4 | < 0.001 |
| Male | 97.5% | 83.4% | <0.001 |
| White | 72.5% | 63.1% | <0.001 |
| Hispanic | 7.0% | 9.1% | <0.001 |
| O2 supplementation | 1.8% | 0.7% | <0.001 |
| Current smoker | 8.9% | 12.0% | <0.001 |
| Charl2Yrs | 3.23 ± 2.65 | 1.41 ± 2.00 | <0.001* |
| CharlEver | 4.99 ± 3.31 | 2.28 ± 2.77 | <0.001* |
| Elix2Yrs | 11.87 ± 12.75 | 4.77 ± 8.45 | <0.001* |
| ElixEver | 21.28 ± 16.62 | 9.47 ± 12.76 | <0.001* |
| PDeathDx | 0.139 ± 0.128 | 0.049 ± 0.064 | <0.001* |
| PDeathLabs | 0.136 ± 0.106 | 0.058 ± 0.065 | <0.001* |
| AggRiskRx | -0.0854 ± 0.2720 | -0.2025 ± 0.5424 | <0.001* |

\* Mann-Whitney U-test; differences in ages were tested by student's t-test

The main multivariate model is shown in **Table 2**. 239,393 patients (70.5%) had complete data sets available for multivariate modeling. 13 variables were identified as statistically significant and independent determinants of death at 60 days. A poorer prognosis was observed for the elderly, males, and Hispanics while being white was protective. PDeathDx, PDeathLabs, AggRiskRx, and 3 of 4 comorbidity measures were all significant predictors of death. The adjusted odds ratio for delta infection was 1.87 ± 0.05 which corresponds to a relative risk of 1.78. The adjusted odds ratio for prior vaccination was 0.280 ± 0.011 which corresponds to a relative risk of 0.291. This observation suggests that the delta variant is substantially more lethal than earlier variants – an effect that is largely offset by prior vaccination.

**TABLE 2:**
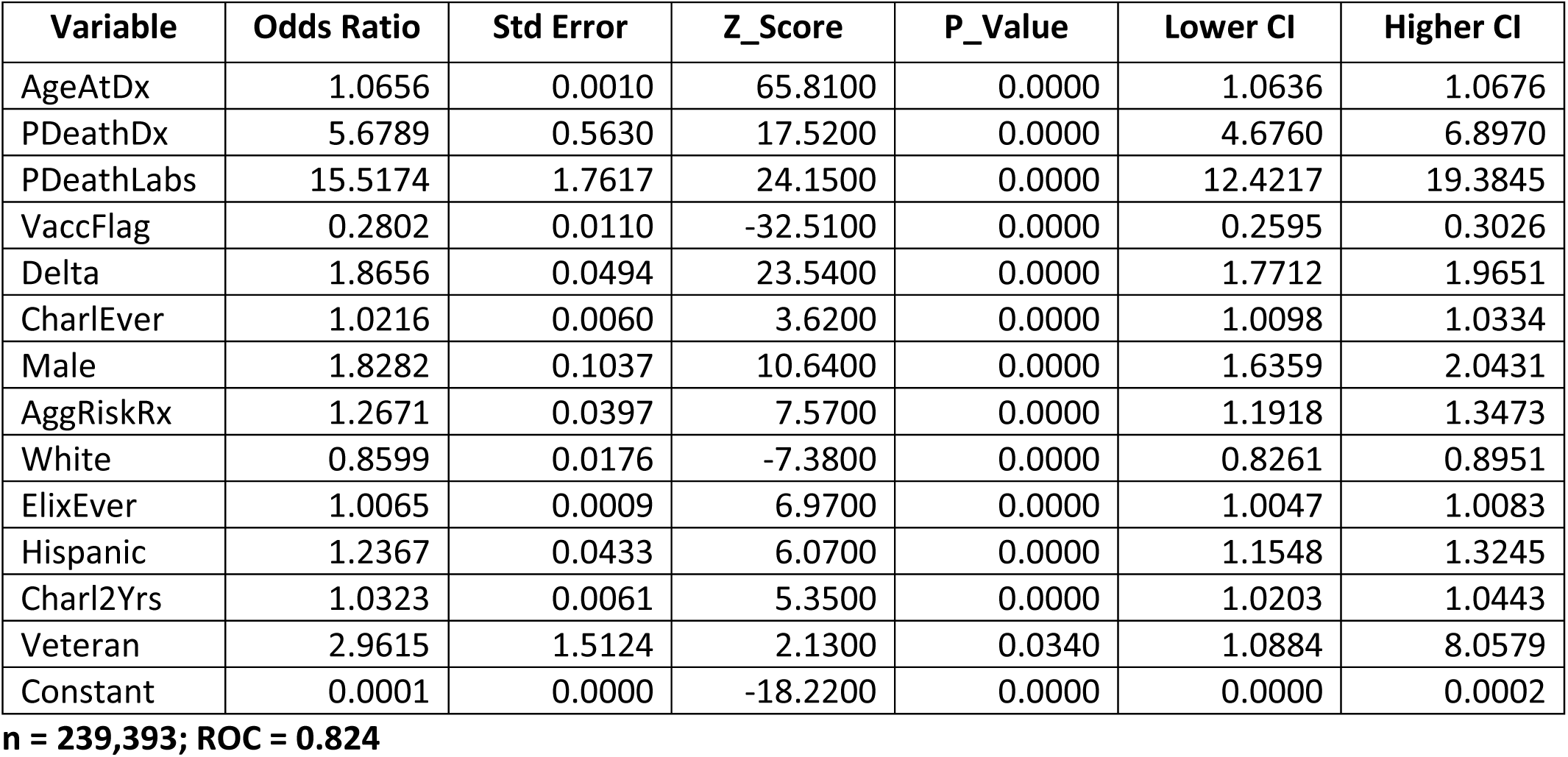
MAIN MULTIVARIATE MODEL.

**Tables 3** and **4** show the multivariate models for early COVID variants and delta, respectively. Of 11 variables identified as predictors before July 1, 2021, 8 were still significant after the emergence of delta. The adjusted odds ratio for vaccination prior to July 2021 was 0.404 ± 0.033 while the odds ratio thereafter was 0.259 ± 0.012. This observation suggests that prior vaccination was more effective in reducing the case fatality rate for delta than earlier variants. However, only 4,649 (or 15.1%) of 18,120 breakthrough infections occurred before July 1. The earlier odds ratios were therefore based on a relatively small number of deaths in the vaccinated group.

**TABLE 3:** MULTIVARIATE MODEL FOR EARLY VARIANTS.

| Variable | Odds Ratio | Std Error | Z_Score | P_Value | Lower CI | Higher CI |
| --- | --- | --- | --- | --- | --- | --- |
| AgeAtDx | 1.0681 | 0.0012 | 60.0600 | 0.0000 | 1.0658 | 1.0704 |
| PDeathDx | 5.7727 | 0.6256 | 16.1800 | 0.0000 | 4.6680 | 7.1388 |
| PDeathLabs | 14.6853 | 1.8320 | 21.5400 | 0.0000 | 11.5000 | 18.7530 |
| CharlEver | 1.0255 | 0.0067 | 3.8400 | 0.0000 | 1.0124 | 1.0388 |
| VaccFlag | 0.4039 | 0.0332 | -11.0400 | 0.0000 | 0.3438 | 0.4744 |
| Male | 1.8551 | 0.1233 | 9.2900 | 0.0000 | 1.6285 | 2.1133 |
| White | 0.8241 | 0.0187 | -8.5500 | 0.0000 | 0.7883 | 0.8614 |
| AggRiskRx | 1.3161 | 0.0485 | 7.4500 | 0.0000 | 1.2244 | 1.4147 |
| Hispanic | 1.2593 | 0.0488 | 5.9500 | 0.0000 | 1.1672 | 1.3585 |
| ElixEver | 1.0059 | 0.0010 | 5.7100 | 0.0000 | 1.0039 | 1.0080 |
| Charl2Yrs | 1.0250 | 0.0067 | 3.7600 | 0.0000 | 1.0119 | 1.0383 |
| Constant | 0.0002 | 0.0000 | -87.3900 | 0.0000 | 0.0002 | 0.0003 |

n = 186,505; ROC = 0.826

**TABLE 4:** MULTIVARIATE MODEL FOR DELTA VARIANT.

| Variable | Odds Ratio | Std Error | Z_Score | P_Value | Lower CI | Higher CI |
| --- | --- | --- | --- | --- | --- | --- |
| AgeAtDx | 1.0570 | 0.0021 | 27.5400 | 0.0000 | 1.0529 | 1.0612 |
| VaccFlag | 0.2593 | 0.0119 | -29.4700 | 0.0000 | 0.2370 | 0.2836 |
| Charl2Yrs | 1.0704 | 0.0119 | 6.1100 | 0.0000 | 1.0473 | 1.0941 |
| PDeathLabs | 20.0041 | 5.5087 | 10.8800 | 0.0000 | 11.6604 | 34.3181 |
| PDeathDx | 4.5774 | 1.1316 | 6.1500 | 0.0000 | 2.8196 | 7.4311 |
| Male | 1.9115 | 0.2051 | 6.0400 | 0.0000 | 1.5490 | 2.3589 |
| ElixEver | 1.0094 | 0.0018 | 5.2800 | 0.0000 | 1.0059 | 1.0129 |
| O2Supp | 1.8408 | 0.2729 | 4.1200 | 0.0000 | 1.3766 | 2.4616 |
| AggRiskRx | 1.1373 | 0.0664 | 2.2000 | 0.0270 | 1.0144 | 1.2751 |
| Constant | 0.0007 | 0.0001 | -46.2300 | 0.0000 | 0.0005 | 0.0010 |

n = 52,888; ROC = 0.818

73,117 patients were presumed to have been infected with the delta variant based their date of infection. 26,168 (35.8%) had previously been vaccinated. 25,818 vaccinees were assigned to 8 cohorts defined by the time from completed vaccination to diagnosis (VxToDx) (in 4-week blocks). Cohort 1 was comprised of the most recent vaccinees while cohort 8 had the most remote vaccinations. The cohorts varied in size from 457 to 6,896 subjects. **Figure 1** shows that CFR fell across the lowest cohorts and reached a nadir of 2.19% for cohort 3. It then increased monotonically across cohorts 4 to 8. One possibility for the latter trend is that vaccine effectiveness declined after 10-14 weeks. However, patients in the later cohorts (4-8) also received their vaccinations the earliest because their needs were the most urgent. For this reason, we used a dual approach to control for confounding. 52,613 patients (72.0% of all delta cases) were available for this analysis. For each cohort of interest, patients in the other cohorts were excluded and a common model fitted to the remaining cases. Thus, members of each cohort had the same priority for vaccination, and the effect of vaccination for each cohort was adjusted for 8 other patient attributes. **Figure 2** shows that the adjusted odds ratio for vaccination declined across the lowest cohorts and remained low for the remaining ones. Patients with the most remote vaccinations still had an adjusted odds ratio for vaccination of 0.244 ± 0.012. Thus, there was no evidence that the vaccine effect on CFR declined over the observation period in this study.

## DISCUSSION

In this paper, we stress the importance of a robust system for classifying vaccine effects. The reason is that the usual methods for evaluating effectiveness are composite measures reflecting 3 underlying risks (exposure, infection, death) and may not precisely define the mechanisms by which vaccines have failed. For example, the mortality rate is a function of the probabilities of exposure, developing an infection once exposed, and dying once infected. Improvements in the last 2 risks (i.e., for infection and death) may be offset by increases in the first (i.e., exposure) as patients abandon COVID precautions. On the other hand, CFR (case fatality rate) is a function of only the risk of death once infected. In this study, we determined the extent to which vaccination provides protection against mortality using a metric not affected by masking, social distancing, handwashing, early testing, and quarantines.

Observational studies of vaccine effectiveness are heavily biased. Patients at the highest risk of death are more likely to receive the vaccine for several reasons including personal choice, concern of their physicians, and/or national policies driven by vaccine shortages and stressed delivery systems. This prioritization confounds the relationship between the intervention and outcome because the benefits of vaccination are offset by their preferential use in patients with the poorest prognosis. The accuracy of estimates for vaccine effect depends upon the extent to which this bias is removed. Our study is unique in that we performed a systematic review of major domains in the medical record, identified observations associated with a poor outcome, and used summary measures of the findings to adjust the effects of vaccination. The review included clinical measurements not tested as candidate variables in other models but are critical determinants of survival (such as oxygen saturation). This approach is innovative because it represents intensive computer processing of hundreds of millions of observations to identify hundreds of potential confounders.

We found that the adjusted odds ratio for vaccination was 0.280. This value corresponds to a 71% reduction in the risk of death. This benefit was observed at a median of 5.1 months after vaccination. Substantial benefits of vaccination were observed before and after the emergence of delta, although the former was significantly less.

Our cohort studies showed that CFR for vaccinees declined with time to a nadir 10-14 weeks after vaccination and then rose thereafter. Since CFR is not affected by patient behaviors or environmental factors, this pattern is consistent with the acquisition and subsequent loss of a physiological factor that promoted recovery from an established infection. The other possibility is a strong selection bias. Patients with the longest time from vaccination to diagnosis received their vaccines the earliest. For example, for delta infections acquired in August 2021, cohort 8 contained the very first vaccinees while cohort 1 contained patients who deferred their vaccinations until July. Because early vaccinations were directed at those at highest risk of death, CFR would be higher for early vaccinees regardless of vaccine effect. For this reason, we used a dual approach to confounders to control for the urgency of vaccination as well as other patient attributes. Our results showed that the rising CFR with time was due to selection bias and not loss of vaccine effect. The benefits of vaccination remained large even at 30-34 weeks – the longest observation period in this study. This finding contrasts sharply with prior studies (1-3) showing a loss of vaccine effect over time. The difference may be due to the use of CFR instead of infection or mortality rates, more robust handling of confounders, and avoidance of selection biases introduced by the way that VA rolled out its vaccination program. Unlike infection or mortality rates, CFR is not affected by unmeasured personal behaviors or environmental factors that affect the probability of exposure. The odds ratios for vaccination in our models were also adjusted for a much larger number of pre-existing conditions, vital sign abnormalities, laboratory results, and medications than previously reported. Finally, because our cohorts were assembled over a short-time frame, our approach was not affected by VA’s changing priorities for vaccination.

We do not have an explanation for why it took 10-14 weeks for vaccination to reach its maximal effect. Future studies should be done on the mechanisms by which COVID vaccination produces such late effects. Regardless of the reasons, our study suggests that COVID precautions should be observed for several weeks after vaccination until the full benefits are attained.

Prior vaccination has been included in other prediction models for COVID mortality. For example, Hippisley-Cox et al (8) published a multivariate model for COVID death in a large, vaccinated cohort in England. The investigators found that the adjusted hazard ratio for full versus partial vaccination was 0.17 (0.13 to 0.22), suggesting a major effect. Differences in vaccine effectiveness between our study and theirs may be explained by the populations studied. To be eligible for military service (and thus inclusion in the present study), individuals cannot have a debilitating congenital abnormality. It is also possible that veterans may have diminished immune responses to vaccination that may not be fully explained by covariates even in a highly specified model. The effect of vaccination on the case fatality rate may therefore depend upon the population studied.

Our definition of a delta infection relied on information provided by the Centers for Disease Control (CDC). CDC identifies and tracks COVID variants through genomic surveillance. It collects specimens for sequencing through the National SARS-CoV-2 Strain Surveillance program as well as data from commercial and academic laboratories under contract to state and local public health agencies. *Nowcast* is their model for reporting proportions of the variants by region and week. CDC announced that delta became the dominant variant on July 7 and that the variant accounted for 83% of isolates by July 20. These findings corresponded to an alarming increase in the 7-day moving average for reported cases from 12,000 in late June to over 60,000 by July 27, 2021. This increase prompted CDC to issue new guidance on vaccinations and personal precautions for delta on that date. For the week of October 4, 2021, delta accounted for 99.2% of reported cases – a date which corresponds to the end of our sampling period. It was not overtaken by omicron as the dominant variant until the week of December 19, 2021. These data suggest that the great majority of cases presenting between July - September 2021 had delta. Our study showed that these patients had a worse prognosis compared to preceding cases and that the difference could not be explained by patient attributes or vaccination. A different approach was used by groups studying delta in England and Canada (9, 10). These groups studied patients with variants confirmed by viral genome sequencing and likewise found a poor prognosis for delta. Another difference between our study and theirs relates to the control of confounders. Poor outcomes can be attributed to the virulence of the variant, the degree to which the host response is compromised by pre-existing conditions, or both. Measures of virulence are valid to the extent that the latter is controlled. Although outcomes in their models were adjusted for demographic characteristics, one model (9) did not include comorbidities while the other included the presence of “any documented major comorbidity” (10). Our novel approach to pre-existing conditions, vital signs, laboratory tests, and outpatient medications enable more rigorous control of cofounders than previously reported methods. In fact, we could not demonstrate that delta had a poorer outcome until these factors and vaccination status were controlled. This issue becomes more important as each surge culls the most vulnerable patients in a population – leading to a gradual change of patient characteristics in the susceptible population. Although they differ in design, all 3 studies confirm that the outcomes of delta infection are poorer and that the variant is more virulent than early variants.

Our models did not include the use of anti-virals, dexamethasone, anticoagulation, or monoclonal antibodies. The reason for such is that the most commonly used anti-viral agent – remdesivir – has little effect on patient mortality (11, 12). In addition, the effectiveness of dexamethasone treatment is not definitive, guidelines for anticoagulation therapies have not been fully formulated, and the use of monoclonal antibodies remains typically restricted. However, as effective anti-virals and other therapies are utilized on future cases, our models will be modified to evaluate their effects.

Although our findings show the effectiveness of vaccination, one cannot definitively prove that vaccination improved survival. For example, while we controlled for as many clinical variables as possible, there is no way of measuring all relevant patient attributes. Vaccination could be a marker for many traits that affect recovery from serious illness such as physical fitness, nutrition, medication compliance, preventive care and so forth. This possibility is suggested by the observation that vaccinated patients have a lower risk of death than unvaccinated persons even when COVID deaths are excluded (13).

Our conclusions are limited to patients with characteristics of the veteran population. Because our approach requires an extensive amount of baseline data, the results are also biased towards patients with chronic conditions that require periodic evaluation. Our methods represent a new approach to evaluating the effectiveness of interventions in observational studies. If validated by others for COVID and other diseases, the approaches presented here represent an alternative and perhaps more robust method for reconciling confounders.

## Data Availability

All data in the present study are available upon reasonable request to the authors

## Notes

### Competing Interest Statement

The authors have declared no competing interest.

### Funding Statement

This study did not receive any funding

### Author Declarations

Institutional Review Board of the New Mexico VA Health Care System gave ethical approval for this work

### Summary of Updates

The title has been revised to be more informative. The introduction was re-written to clarify the aims and study design.

